# News Coverage and Drug Shortages during the COVID-19 Pandemic

**DOI:** 10.1101/2020.10.12.20211656

**Authors:** Maryaline Catillon, Maimuna S. Majumder, Shannon F. Manzi, Kenneth D. Mandl

## Abstract

Several drugs repurposed as COVID-19 treatment are in short supply. We collect data from MediaCloud and Google Health Trends about eight drugs proposed for repurposing as COVID-19 treatments and reported to be in shortage by the U.S. Food and Drug Administration (FDA) from January 1, 2020 through June 30, 2020. We find that news media coverage could have contributed to shortages due to hoarding by individuals and stockpiling by institutions, and that search trends appear to accurately discriminate between individual hoarding and institutional stockpiling.

## Background

In recent months, multiple drugs have been proposed for repurposing as COVID-19 treatments (DPRCT),^1,2^ often with little evidence. Several of these drugs have come into national shortage. We assessed whether drug hoarding by individuals and drug stockpiling by institutions could be associated with media coverage.

## Objective

We sought to analyze U.S. news coverage and Internet search volume for eight DPRCT reported to be in shortage by the U.S. Food and Drug Administration (FDA) from January 1, 2020 through June 30, 2020.

## Methods

DPRCT were selected and categorized by SM, a pharmacist involved in federal COVID response, based on information from the American Society of Hospital Pharmacists (https://www.ashp.org/Drug-Shortages/Current-Shortages) and national and international workgroups. Because patients can directly purchase these medications, three DPRCT were characterized as able to be hoarded by individuals or stockpiled by institutions: azithromycin, famotidine and hydroxychloroquine. Five DPRCT, which are not generally purchased directly by patients, were characterized as able to be stockpiled by institutions only: cisatracurium, dexmedetomidine, continuous renal replacement therapy (CRRT), midazolam and propofol.

For each DPRCT, U.S. news volume was measured with MediaCloud’s National Corpus (https://sources.mediacloud.org/#/collections/34412234) and reported separately for (1) news about the drug excluding mention of the word “shortage” and (2) news about the drug including the word “shortage”. U.S. Internet search volume was ascertained using Google Health Trends, with queries for each drug name. The shortage date announcements were based on reports in the FDA Drug Shortages Database (https://www.accessdata.fda.gov/scripts/drugshortages/default.cfm).

## Results

For all DPRCT that might be stockpiled by institutions or hoarded by individuals (Figure 1), news about the drug excluding the word “shortage” and news about the drug shortage both spike before the shortage announcement. For all three drugs, Internet searches also spike before the shortage announcement.

**Figure 1.**
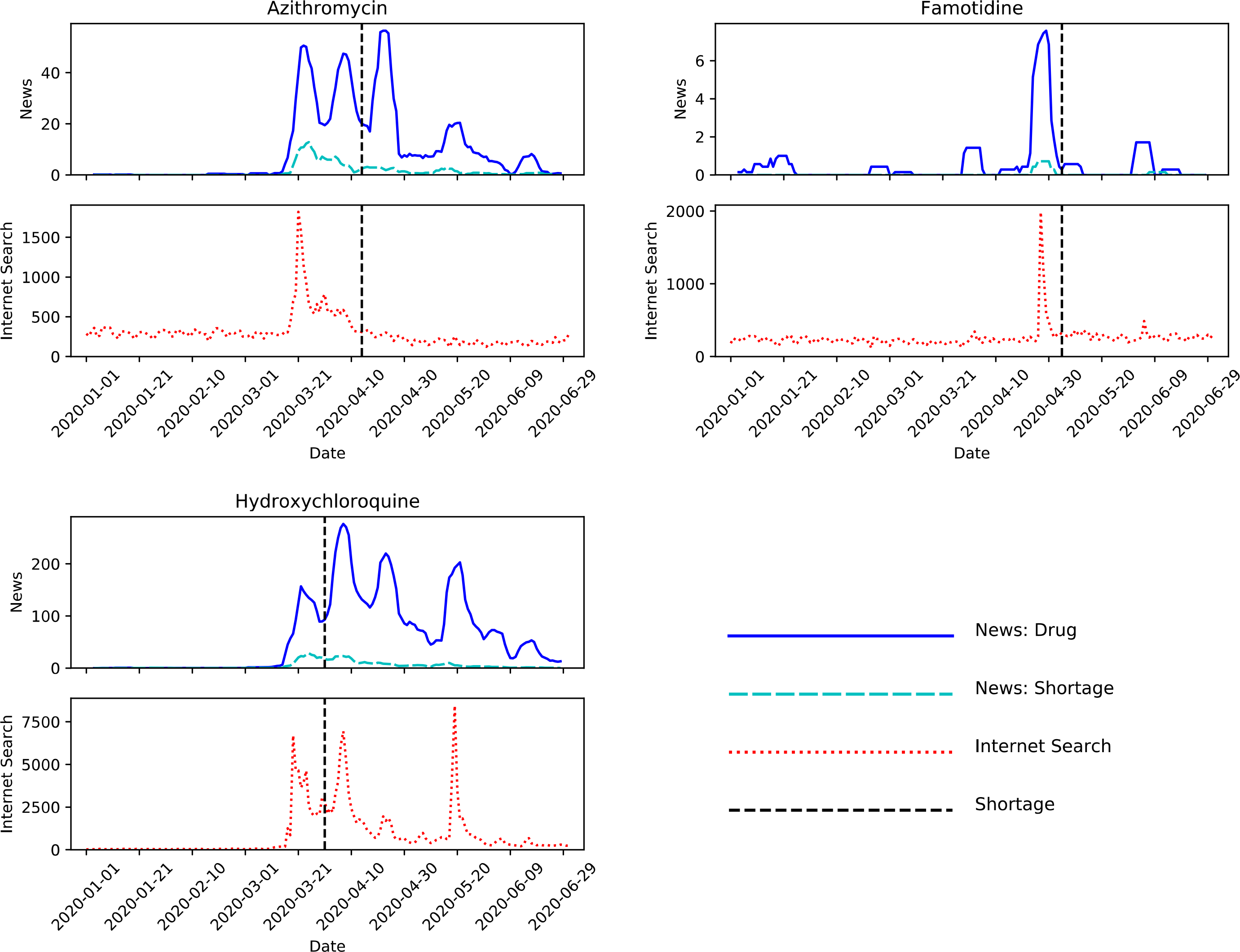
News volume, Internet search trends, and shortage date for azithromycin, famotidine and hydroxychloroquine. News volume (7-day rolling average), from MediaCloud’s U.S. National Corpus is reported separately for (1) news about the drug excluding mention of the word “shortage”, labelled “News: Drug” and (2) news about the drug including the word “shortage”, labelled “News: Shortage.” Internet search volume was measured using Google Health Trends, with queries for each drug name. Shortage date announcements are from the Food and Drug Administration (FDA) Drug Shortages Database.

For four of five DPRCT which might be stockpiled by institutions but are not ordinarily hoarded by individuals (Figure 2), news about the drug excluding the word “shortage” and news about the drug shortage also spike before the announcement, but for all five drugs Internet search trends are flat.

**Figure 2.**
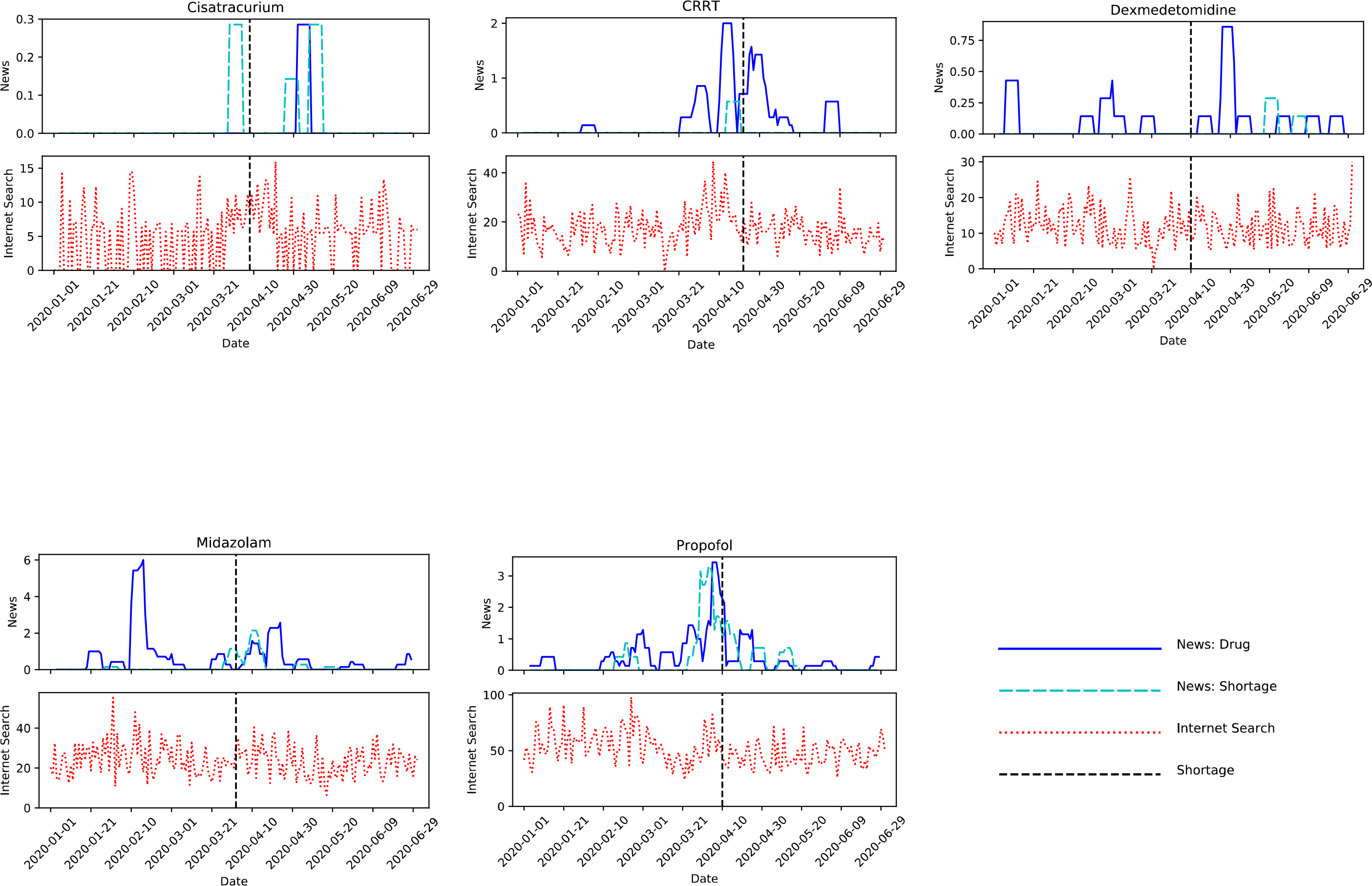
News volume, Internet search trends and shortage date for Cisatracurium, Continuous renal replacement therapy, dexmedetomidine, midazolam, and propofol. News volume (7-day rolling average), from MediaCloud’s U.S. National Corpus, is reported separately for (1) news about the drug excluding mention of the word “shortage”, labelled “News: Drug” and (2) news about the drug including the word “shortage”, labelled “News: Shortage.” Internet search volume was measured using Google Health Trends, with queries for each drug name. Shortage date announcements are from the Food and Drug Administration (FDA) Drug Shortages Database.

## Discussion

Though temporality does not establish causality, all but one of the DRPCT shortages were preceded by spikes in news reports, consistent with an impact of news on purchasing behaviors.^3^ Media coverage may trigger stockpiling by institutions and hoarding by patients. Such behaviors risk depriving patients who are dependent on those drugs for approved and widely-accepted off label uses.

Spikes in Internet searches, previously demonstrated as a potential proxy for medication purchase intent,^5,6^ suggest that news influenced individuals to research and acquire DPRCT that they can purchase directly. In contrast, DRPCT that could only be stockpiled by institutions did not exhibit search spikes prior to shortages. Because we don’t generally expect institutions to conduct Internet searches prior to purchasing drugs, search trends appear to accurately discriminate between individual hoarding and institutional stockpiling.

## Data Availability

The data is available from MediaCloud and Google Health Trends.

## Data Availability

The data is available from MediaCloud and Google Health Trends.

## Notes

### Competing Interest Statement

Maryaline Catillon is an Associate at Analysis Group. The opinions expressed are those of the authors and do not necessarily reflect the views of the Analysis Group, Inc., its clients, or any of its or their respective affiliates.

### Funding Statement

No external funding was received.

